# Race, Ethnicity and Their Implication on Bias in Large Language Models

**DOI:** 10.64898/2026.01.04.26343415

**Authors:** Shiyue Hu, Ruizhe Li, Yanjun Gao

## Abstract

Large language models (LLMs) increasingly operate in high-stakes settings including healthcare and medicine, where demographic attributes such as race and ethnicity may be explicitly stated or implicitly inferred from text. However, existing studies primarily document outcome-level disparities, offering limited insight into internal mechanisms underlying these effects. We present a mechanistic study of how race and ethnicity are represented and operationalized within LLMs. Using two publicly available datasets spanning toxicity-related generation and clinical narrative understanding tasks, we analyze three open-source models with a reproducible interpretability pipeline combining probing, neuron-level attribution, and targeted intervention. We find that demographic information is distributed across internal units with substantial cross-model variation. Although some units encode sensitive or stereotype-related associations from pretraining, identical demographic cues can induce qualitatively different behaviors. Interventions suppressing such neurons reduce bias but leave substantial residual effects, suggesting behavioral rather than representational change and motivating more systematic mitigation.

## 1 Introduction

Large language models (LLMs) are increasingly used in high-stakes domains such as healthcare, where demographic attributes (e.g., race, ethnicity, gender) may be explicitly stated or implicitly inferred from text. Prior work shows that LLMs can condition their outputs on demographic information even when it is not task-relevant (Zack et al., 2024; Kim et al., 2023; Fraser and Kiritchenko, 2024; Zhao et al., 2025), therefore can induce mis-attribution on model output with undesirable or biased behavior (Demchak et al., 2024; Levartovsky et al., 2025; Zack et al., 2024).

Most prior studies on demographic bias focus on outcome-level effects, evaluating disparities in generated responses, accuracy, calibration, or toxicity scores across demographic groups (Tan and Lee, 2025; Hartvigsen et al., 2022; Guan et al., 2025; Wang et al., 2025). While these analyses are essential for documenting harm, they treat LLMs as black boxes, offering limited insight into whether demographic attributes are encoded as high-level semantic features, task-relevant representations, or spurious shortcuts during prediction. In parallel, recent works in mechanistic interpretability demonstrated how LLMs encode demographic information and manipulated internal LLMs’ states to ensure fairness (Yu and Ananiadou, 2025; Ahsan and Wallace, 2025; Karvonen and Marks, 2025), yet these tools have rarely been applied to demographic bias in a systematic and task-diverse manner.

A central challenge is that demographic attributes interact with language in complex ways. In many real-world settings, demographic information may be explicitly stated (e.g., “a Black patient,” “a Hispanic speaker”) or implicitly conveyed through linguistic, cultural, or geographical cues, i.e. the “proxy” cues. Moreover, the same demographic signal can have qualitatively different effects depending on the task: it may alter predicted medical risk in a clinical scenario, while simultaneously modulating perceived toxicity, credibility, or intent in open-ended generation. Existing evaluation typically isolate a single task or domain (Hartvigsen et al., 2022; Zack et al., 2024; Levartovsky et al., 2025), making it difficult to assess whether demographic sensitivity reflects general representational mechanisms or task-specific heuristics.

In this work, we investigate how demographic attributes influence LLM behavior, with a focus on mechanistic explanations rather than surface-level disparities. We examine *race* and *ethnicity* as commonly occurring coarse-grained categories (e.g. White, Black, Asian, Hispanic and Latino) as they appear in the studied datasets, rather than attempting to model the full sociological complexity of these constructs. Using two publicly available datasets, we study: 1) toxicity-related generation tasks (Hartvigsen et al., 2022), where the same attributes may alter the likelihood, tone, or framing of model outputs, and 2) clinical narrative tasks (Bear Don’t Walk IV et al., 2024), where the same attributes appear through explicit or indirect cues in medical text and modulate model behavior despite identical clinical evidence.

We adopt a mechanistic interpretability (MI) framework to study how lexical cues of race and ethnicity are encoded and propagated within three open-source LLMs that are widely used: Qwen2.5-7B (Team, 2024), Mistral-7B (Jiang et al., 2023), and Llama-3.1-8B (Grattafiori et al., 2024). Our contributions are threefold:

- a reproducible MI **pipeline** that combines multi-class probing, neuron-level attribution, and targeted intervention to identify internal units associated with demographic attributes and to examine their functional relevance across tasks. The proposed framework is applicable to other social variables beyond race and ethnicity.
- a fine-grained **characterization** of race and ethnicity representations across LLMs, revealing the distributed nature of demographic information and model-specific emphasis on semantic facets such as geography, language, culture, or historical context.
- a mechanistic **analysis** of how demographic representation influences model behaviors. Although internal features encode sensitive or harmful stereotype-related concepts present in pretraining data, these representations are unevenly activated by direct and indirect demographic cues.

Our findings show that while race- and ethnicity-associated representations can be identified and analyzed at the neuron level, their associations with biased model behavior persist even when highly active neurons are surpressed. This indicates that biased behavior in LLMs cannot be fully explained or controlled by manipulating a small set of identifiable neurons alone.

## 2 Related Work

### Mechanistic Interpretability of Bias in LLMs

Recent works in mechanistic interpretability have begun to locate where demographic information is encoded. Our approach of using probe-based neuron localization aligns with emerging research in this space. Yu and Ananiadou (2025) utilized neuron editing to understand and mitigate gender bias, identifying specific “gender neurons” within the MLP layers. In the medical domain, Ahsan et al. (2025) investigated the mechanisms of demographic bias specifically for healthcare tasks, suggesting that certain internal representations are disproportionately sensitive to racial identifiers. More recently, Ahsan and Wallace (2025) explored the use of Sparse Autoencoders (SAEs) to reveal clinical racial biases. Our work extends these findings by demonstrating that race-specific neurons are not only present in general datasets but are consistently activated and influential on domain-specific text.

### Internal Bias Mitigation and Steering within LLMs

Beyond identification, recent work focuses on manipulating internal model states to ensure fairness. Zhou et al. (2024) proposed the UniBias framework, which mitigates bias by manipulating attention heads and MLP components. For real-time applications, Li et al. (2025) introduced *FairSteer*, a dynamic activation steering method that adjusts model behavior during inference. Karvonen and Marks (2025) further demonstrated that interpretability-based interventions can improve fairness more robustly than traditional fine-tuning in realistic settings. Our methodology contributes to this line of work by providing a targeted intervention strategy, specifically sign-flipping and amplification, to suppress biased pathways. This builds upon the “context-aware” fairness frameworks suggested by Nadeem et al. (2025), ensuring that mitigation is grounded in the semantic understanding of the racial directions we extract.

### Racial Bias in Clinical LLMs

Extensive research has documented that LLMs inherit and propagate racial biases when applied to clinical decision support. Zhang et al. (2023) demonstrated that ChatGPT exhibits disparate treatment recommendations for acute coronary syndrome based on racial and gender cues. Similarly, Zack et al. (2024) evaluated GPT-4, finding that model frequently perpetuates harmful stereotypes that could lead to inequitable health outcomes. Poulain et al. (2024) further expanded this analysis across various clinical decision-support tasks, highlighting that bias patterns are not idiosyncratic but systematic across model families. While these studies establish the existence of bias, they largely treat model as a black box. Our work seeks to uncover the internal mechanisms driving these disparate outputs.

## 3 Background

### MLP Layers and Neuron Activation

Modern Transformer-based LLMs process information through a *residual stream*. In this framework, the residual stream acts as a communication channel, while MLP layers function as key-value memories that store and inject factual associations into the stream (Geva et al., 2021a). Contemporary models like Llama 3.1, Mistral, and Qwen 2.5 utilize the *SwiGLU* gated architecture (Shazeer, 2020). The output of an MLP block with input *x* is defined as:

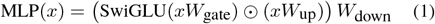

where ⊙ is the element-wise product. We define an individual *neuron j* as the *j*-th element of the intermediate gated state. The total MLP output is the sum of these neurons’ contributions:

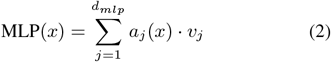

where *a*_*j*_(*x*) is the activation score (the product of the gate and up-projections) and *v*_*j*_ is the *j*-th row of *W*_down_. Our method specifically probes these *output vectors v*_*j*_ to locate racial information.

### Logit Lens

To interpret high-dimensional vectors in residual stream or neuron output vectors *v*_*j*_, we use *Logit Lens* (nostalgebraist, 2020). This technique projects a vector *h* directly into vocabulary space using model’s unembedding matrix *W*_*U*_: logits = *hW*_*U*_. By inspecting top-ranked tokens in the resulting distribution, we can decode the semantic concepts encoded within specific neurons.

## 4 Methodology

We propose a mechanistic interpretability framework to determine *where* and *how* race information is encoded within LLMs. Our approach progresses from identifying global race directions via multiclass probing to identifying the specific neurons responsible for these encodings.

### 4.1 Locating Race Directions via Multi-Class Probing

To extract race representations, we train linear probes *W*_Race_ to classify racial group membership for each model. The probe is trained on the final-layer residual stream 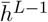, averaged across all token positions:

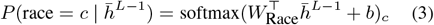

where *W*_Race_ ∈ ℝ^*d×*|𝒞|^ is the learned probe matrix, *b* is bias vector, and 𝒞 denotes the set of racial super-categories (e.g., Asian, Black, Middle Eastern). Each column *w*_*c*_ of *W*_Race_ represents *race direction* for group *c* in model’s representation space.

### 4.2 From Race Directions to Neurons

Having identified the global race directions *w*_*c*_, we locate the MLP neurons that write to these directions, motivated by prior work showing MLPs act as key – value memories (Geva et al., 2021b).

#### Interpreting the Probe Direction

We first verify that our learned directions *w*_*c*_ capture meaningful racial semantics. Using Logit Lens, we project each direction into vocabulary space via the model’s unembedding matrix *W*_*U*_:

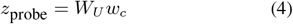

The top-*k* tokens (we use *k* = 20) with the highest values in *z*_probe_ serve as a semantic fingerprint for each racial group.

#### Identifying Candidate Neurons

To locate neurons that write to race direction, we compute the cosine similarity between each MLP neuron’s output vector 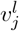 at layer *l* and the probe direction *w*_*c*_:

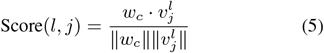

All neurons in final four MLP layers are ranked by this score, and the top 20 candidates are selected. Each candidate is selected by projecting its output vector into vocabulary space and inspecting top-20 tokens. Neurons are retained only if their tokens show clear alignment with the target racial group.

### 4.3 Validating Neurons via Activation Analysis and Intervention

To confirm that identified neurons encode meaningful racial information and causally influence model behavior, we design a two-stage validation pipeline.

#### Activation Analysis

We measure how strongly each neuron group activates across different inputs. For a given input text, we extract the activation score of each candidate neuron during the forward pass. For ToxiGen, we average activations across all token positions; for C-REACT, where the model must produce a classification, we extract activations at the final token position where the prediction is made. We then compare activation patterns across racial groups: if neurons identified for a particular race show higher activation scores when processing text about that group compared to other groups, this provides evidence that these neurons selectively respond to race-related content.

#### Targeted Intervention

To test whether identified neurons causally influence model outputs, we manipulate their activations during inference. We implement this using PyTorch forward hooks on the MLP *W*_down_. During the forward pass, we intercept the activation score of each targeted neuron and force it into a strongly negative range. Specifically, we multiply positive activations by a negative factor (e.g., -5) and amplify negative activations by a positive factor (e.g., 5). This ensures that the neuron’s output remains negative, effectively suppressing the encoded feature. This modified signal propagates through the remaining layers, allowing us to measure whether suppressing specific neurons changes model behavior and final outputs.

## 5 Data and Models

### Datasets

We evaluate our framework on two domains to test the generalizability of race encoding mechanisms across different contexts.

ToxiGen (Hartvigsen et al., 2022) is a large-scale machine-generated dataset of toxic statements about minority groups. We use the annotated subset containing 9.9k samples across 13 target groups. Each sample consists of a statement targeting a specific group (e.g., “asians are trying to take over the world with their karate skills” is targeting Asian).

From the 13 target groups, we consolidate related categories into five racial categories: Asian (asian, chinese), Black (black), Latino (latino, mex-ican), Native American (native_american), and Middle Eastern (middle_east, jewish, muslim). We exclude non-racial categories (women, lgbtq, mental_dis, physical_dis).

C-REACT (Contextualized Race and Ethnicity Annotations for Clinical Text (Bear Don’t Walk IV et al., 2024)) provides race and ethnicity annotations for 17,281 sentences drawn from clinical notes in the MIMIC-III database. C-REACT contains real clinical text where race information appears in two forms: direct mentions that explicitly state race (e.g., *“Pt is 42 yo AA female”*) and indirect mentions that imply race through associated attributes such as spoken language or country of origin (e.g., *“Pt required a Spanish interpreter”, “Pt is recently immigrated from France”*). C-REACT provides five racial categories: White, Black/African American (Black/AA), Asian, Native American or Alaska Native, and Native Hawaiian or Other Pacific Islander. However, the dataset is highly imbalanced: zero patients labeled as Native Hawaiian or Other Pacific Islander were found, and only three patients labeled as Native American or Alaska Native. We therefore use three racial categories with sufficient representation: White, Black/AA, and Asian.

### Models

We study three instruction-tuned LLMs of comparable scale from different geographic and cultural training contexts: Llama-3.1-8B-IT (Grattafiori et al., 2024) (US), Mistral-7B-IT-v0.3 (Jiang et al., 2023) (France), and Qwen2.5-7B-IT (Team, 2024) (China). This selection allows us to investigate whether models trained on data from different linguistic and cultural contexts encode racial information differently, given that conceptions of race and ethnicity vary across societies.

## 6 Experiments and Results

### 6.1 Toxigen

Table 1 lists top tokens projected by each race direction. Across models, probes reach similar performance on ToxiGen (around 75% accuracy/macro-F1; Appendix A.1). These tokens capture various facets of racial encoding, including geography, religion, demographic labels, and cultural terms. Across all three models, the learned directions identify tokens that align closely with the target racial groups. This confirms that LLMs store clear racial representations within their residual streams.

**Table 1:**
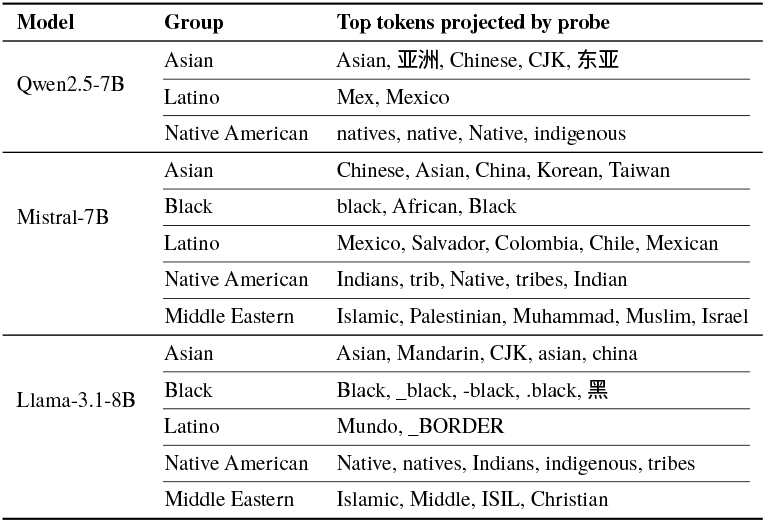
Top tokens by race group across models (ToxiGen). Full results in Appendix A.2. Translations: 亚洲 (Asia), 东亚 (East Asia), 黑(Black).

Table 2 presents race encoding neurons identified within the final four MLP layers. These neurons reveal that LLMs decompose racial concepts into distinct semantic dimensions. Some neurons encode broad demographic terminology that directly names groups, such as Mistral-7B’s 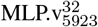 (*Black, black, African*) and Asian neurons across all models (*Asian, Chinese, Japanese*), functioning as explicit demographic classifiers. Others encode race through associated attributes: Llama-3.1-8B’s 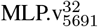 links Asian identity to geographic terms (*Chinese, China, Beijing*), while 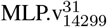 captures Chinese last names (*Li, yuan, Dong, Huang, Wang*); Middle Eastern neurons project to religious and regional identifiers (*Jewish, Judaism, Islam, Jerusalem, Saudi*). We also observe neurons that encode historically harmful associations. Native American neurons across all three models project to colonial terminology (*colonial, colony, colonization*), and neurons for Black identity recover offensive racial terms that persist across models despite different training corpora.

**Table 2:**
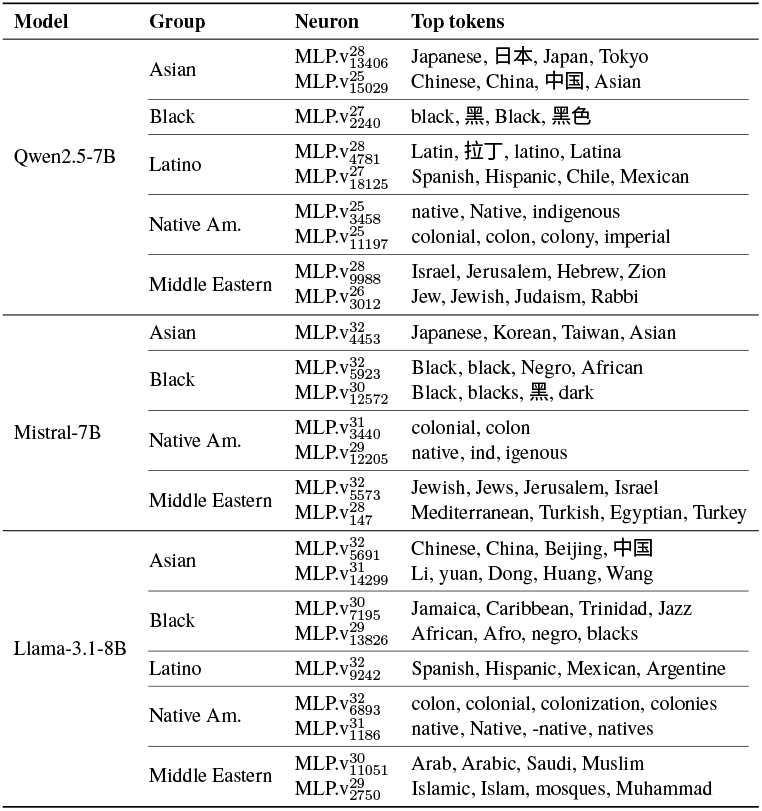
Top race-encoding neurons identified via cosine similarity with probe directions. WARNING: Some tokens reflect harmful stereotypes. Full results in Appendix A.3. Translations: 日本 (Japan), 中国 (China), 黑 (Black), 黑色 (Black color), 拉丁(Latin).

#### Neuron Activation Analysis

To verify that identified neurons selectively respond to their target racial groups, we measure mean activation values when processing test samples from each group. Figure 2 displays these activation patterns as heatmaps, where diagonal cells represent neurons processing their target group. The results confirm that most race encoding neurons activate more strongly for their target group than for others. This is most pronounced for Latino and Middle Eastern neurons: Llama-3.1-8B achieves activation values of 0.83 and 0.90 respectively, while Qwen2.5-7B reaches 0.71 and 1.23. Black neurons also demonstrate consistent selectivity across all three models, with positive diagonal values compared to near zero or negative off diagonal values. Asian and Native American neurons exhibit weaker selectivity, likely reflecting sparser representation in training data. Nevertheless, the overall diagonal pattern validates our neuron identification method: neurons selected via probe direction alignment do preferentially activate for their target groups, validating their role in demographic encoding.

**Figure 1:**
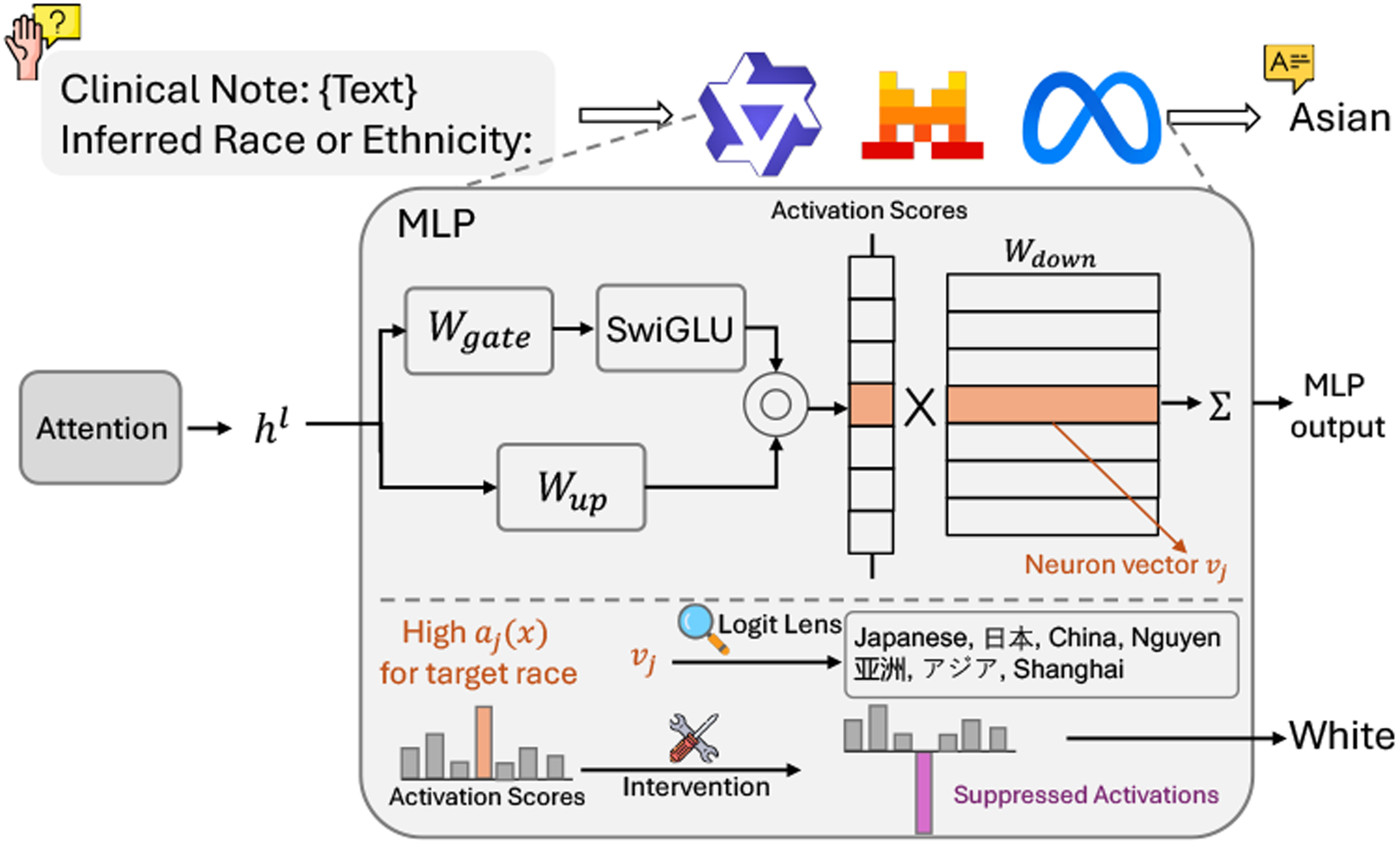
With MLP, we locate neurons relevant to race information and inspect them via Logit Lens. For the higher activation score for target race, we adjust its value to steer model’s behavior.

**Figure 2:**
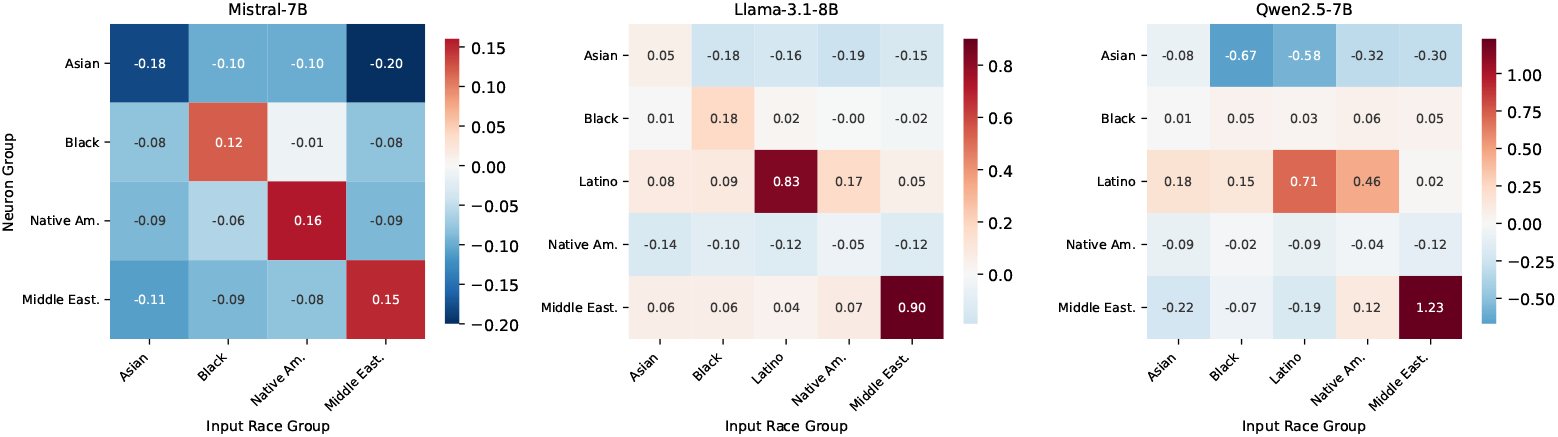
Mean activation values of race encoding neurons when processing text from each racial group (ToxiGen). Diagonal cells represent neurons processing their target group. Higher values (red) indicate stronger activation; lower values (blue) indicate suppression.

### 6.2 C-REACT

We train separate probes on direct and indirect mentions to evaluate whether each type captures distinct representations. Direct-mention probes achieve higher accuracy/F1 than indirect probes (Appendix A.1). This likely reflects that direct cues are explicit and indirect data are sparser. Table 3 compares tokens projected by each probe type. Direct and indirect probes capture semantically distinct representations. Direct probes recover general racial and ethnic terminology (*Asian, African, black*, 白), while indirect probes recover specific countries and regions associated with each group. For instance, White indirect probe projects strongly to Russia and Eastern European terms across all models, reflecting dataset composition where Russian is the most frequent language among White patients. Similarly, Black or African American indirect probes recover Caribbean and African nations (*Haiti, Caribbean, Nigeria*). This divergence confirms that LLMs encode race through multiple pathways: explicit demographic labels and associated geographic or linguistic attributes.

**Table 3:** Comparison of top tokens from direct (race/ethnicity) vs. indirect (language/country) mentions in C-REACT. WARNING: Some tokens reflect harmful stereotypes. Full results in Appendix A.4. Translations: 白(white), 俄罗斯(Russia), 非洲(Africa), 热带(tropical).

| Model | Group | Direct | Indirect |
| --- | --- | --- | --- |
| Qwen2.5-7B | White | 白 | Russian, 俄罗斯, Russia |
|  | Asian | Asian, Asia, Asians, 亚洲 | Chinese, Xia, Tibetan, China |
|  | Black/AA | African, 非洲, Africa, black | Hait, Haiti, Tropical, 热带 |
| Mistral-7B | White |  | Moscow, Russian, Ukrain, Polish |
|  | Asian | Asian, Taiwan, Japanese, Malays | Korea, Korean, Asian, Vietnam |
|  | Black/AA | African, blacks, Negro, slavery | Caribbean, Cuba, Nigeria, Brazil |
| Llama-3.1-8B | White |  | Russia, Kremlin, Putin, Moscow |
|  | Asian | Asian, Indonesian, Asia, Taiwanese | Cambodia, Chinese, wang, Buddhism |
|  | Black/AA | black, African, Afro, negro | Haiti, Caribbean, Dominican, Bahamas |

Tables 4 and 5 present race encoding neurons identified from direct and indirect probes respectively. The neuron projections mirror the probe token patterns: direct mention neurons recover explicit demographic terms (*Asian, Black, African*, 白 *(White)*), while indirect mention neurons recover geographic and cultural associations (*Russia, Moscow, Vietnam, Caribbean*). As in ToxiGen, we observe neurons encoding harmful associations. Qwen2.5-7B’s 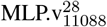 projects to *racist, racism, Harlem, segregation*, and several Black/AA neurons encode terms related to slavery. The persistence of such encodings across both general and clinical domains confirms that these representations are inherited from pretraining rather than induced by domain specific data.

**Table 4:**
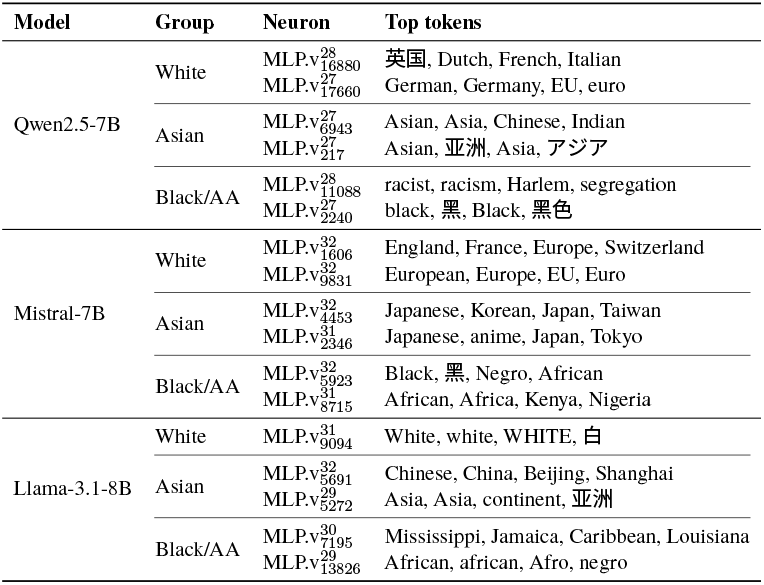
Top race-encoding neurons from C-REACT direct mentions (explicit race/ethnicity). WARNING: Some tokens reflect harmful stereotypes. Full results in Appendix A.5. Translations: 英国 (England), 亚洲 (Asia), アジア (Asia), 黑 (Black), 黑色 (Black color), 白 (White).

**Table 5:**
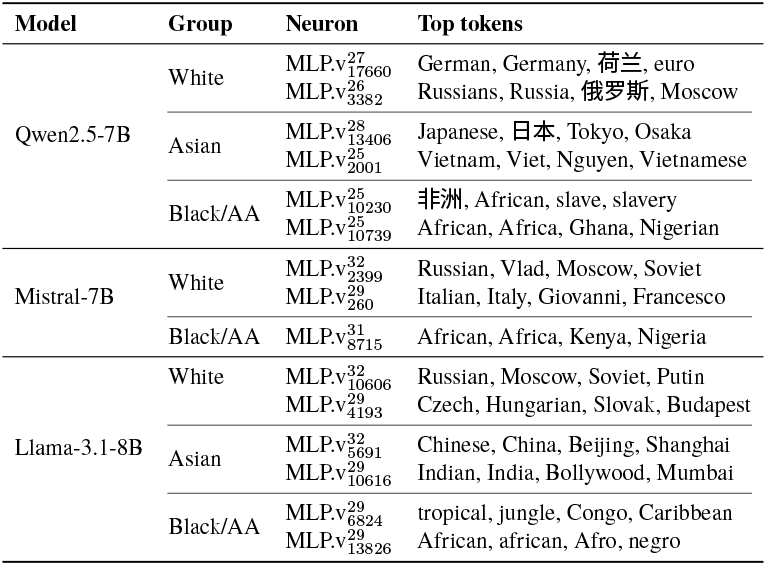
Top race-encoding neurons from C-REACT indirect mentions (language/country). WARNING: Some tokens reflect harmful stereotypes. Full results in Appendix A.6. Translations: 荷兰 (Netherlands), 俄罗斯 (Russia), 日本(Japan), 非洲(Africa).

### 6.3 Neuron Intervention

To test whether the race encoding neurons we identified actually influence model behavior, we design an intervention experiment using C-REACT indirect mentions. Using a template prompt shown in Figure 3, we prompt each model to predict patient race based on clinical text containing only indirect cues such as language or country information, then manipulate race encoding neurons to see if we can correct observed biases.

**Figure 3:**
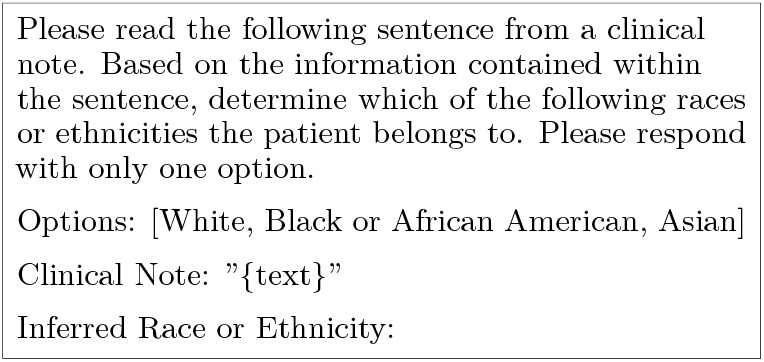
Prompt template for race prediction on C-REACT indirect mentions.

#### Baseline Classification

Table 6 shows misclassification patterns across the three models. The dominant error type varies by model: for Qwen2.5-7B and Llama-3.1-8B, White→Asian misclassification is the primary error, accounting for 75.0% and 95.6% of errors respectively. Llama’s bias is the most pronounced, with 395 of 537 White patients incorrectly classified as Asian. In contrast, Mistral-7B shows a different pattern: its dominant error is White→Black/AA (76.2% of errors). This divergence suggests that models encode and apply racial information differently during inference.

**Table 6:** Misclassification patterns on C-REACT indirect mentions. The dominant error type (bold) varies across models: White→Asian for Qwen and Llama, White→Black/AA for Mistral.

| Error Type | Qwen2.5-7B | Mistral-7B | Llama-3.1-8B |
| --- | --- | --- | --- |
| White→Asian | **27** | 4 | **395** |
| White→Black/AA | 4 | **16** | 5 |
| Black/AA→White | 4 | 0 | 0 |
| Black/AA→Asian | 0 | 0 | 10 |
| Asian→White | 1 | 1 | 1 |
| Asian→Black/AA | 0 | 0 | 2 |
| Total Errors | 36 | 21 | 413 |
| **Dominant Error %** | **75.0%** | **76.2%** | **95.6%** |

#### Activation Patterns

To investigate what drives these biases, we measure activation levels for all neuron groups across all classification outcomes (Table 7). We observe a strong correspondence between neuron groups exhibiting consistently high activation and the dominant error directions identified in Table 6.

**Table 7:** Mean neuron activation scores on C-REACT indirect mention prompts, grouped by classification outcome (*actual* → *predicted*). **W/B/A** denote **White / Black/AA / Asian** (e.g., **W** → **B**: actual White, predicted Black/AA; **W** → **W**: correct). Positive values indicate the neuron group writes in the direction of its output vectors (strengthening the associated signal during generation), while negative values indicate writing in the opposite direction (weakening it). “—” indicates no samples for that outcome.

| Model | Neuron Group | Classification Outcome (Actual → Predicted) |  |  |  |  |  |  |  |  |
| --- | --- | --- | --- | --- | --- | --- | --- | --- | --- | --- |
|  |  | W→W | W→B | W→A | B→W | B→B | B→A | A→W | A→B | A→A |
| Qwen2.5-7B | Asian Direct | **+6.04** | **+6.25** | **+5.85** | **+3.50** | **+5.71** | — | **+4.66** | — | **+3.86** |
|  | Asian Indirect | −1.03 | −1.02 | −0.96 | +0.31 | −0.06 | — | −1.55 | — | −0.33 |
|  | Black/AA Direct | −0.74 | −0.04 | −0.40 | +0.28 | +0.83 | — | +0.27 | — | +0.27 |
|  | Black/AA Indirect | −0.57 | +0.40 | −0.63 | +5.80 | +3.30 | — | −0.39 | — | −0.78 |
|  | White Direct | −3.25 | −3.49 | −3.80 | −2.06 | −3.05 | — | −4.87 | — | −4.01 |
|  | White Indirect | +0.36 | +0.31 | −0.20 | −0.90 | −1.24 | — | −0.33 | — | −1.31 |
| Mistral-7B | Asian Direct | +0.22 | +0.24 | +0.14 | — | −0.15 | — | +0.36 | — | +0.19 |
|  | Black/AA Direct | **+0.82** | **+0.89** | **+0.64** | — | −0.07 | — | **+1.00** | — | **+0.41** |
|  | Black/AA Indirect | −0.56 | −0.42 | −0.55 | — | +0.19 | — | −0.52 | — | −0.44 |
|  | White Direct | +0.10 | −0.06 | −0.02 | — | −0.47 | — | +0.33 | — | +0.37 |
|  | White Indirect | −0.01 | +0.24 | −0.17 | — | −0.27 | — | −0.44 | — | −0.32 |
| Llama-3.1-8B | Asian Direct | −0.53 | −0.59 | −0.59 | — | −0.57 | −0.54 | −0.41 | −0.51 | −0.55 |
|  | Asian Indirect | **+0.27** | **+0.28** | **+0.33** | — | **+0.36** | **+0.43** | **+0.29** | **+0.34** | **+0.47** |
|  | Black/AA Direct | −1.07 | −1.43 | −1.20 | — | −1.97 | −1.66 | −0.54 | −1.60 | −0.96 |
|  | Black/AA Indirect | −0.01 | +0.01 | −0.00 | — | +0.04 | +0.07 | +0.03 | +0.03 | +0.03 |
|  | White Direct | +0.11 | +0.37 | +0.24 | — | +0.51 | +0.49 | +0.62 | +0.34 | +0.70 |
|  | White Indirect | +0.22 | +0.28 | +0.32 | — | +0.01 | +0.04 | −0.04 | +0.01 | −0.01 |

**Table 8:** Test set performance of multi-class linear probes trained on **ToxiGen** (5-way: asian/black/latino/native_american/middle_eastern). We report Accuracy and Macro-F1.

| Model | Acc. (%) | Macro-F1 |
| --- | --- | --- |
| Qwen2.5-7B-IT | 74.88 | 0.74 |
| Llama-3.1-8B-IT | 77.98 | 0.77 |
| Mistral-7B-IT | 75.20 | 0.74 |

**Table 9:**
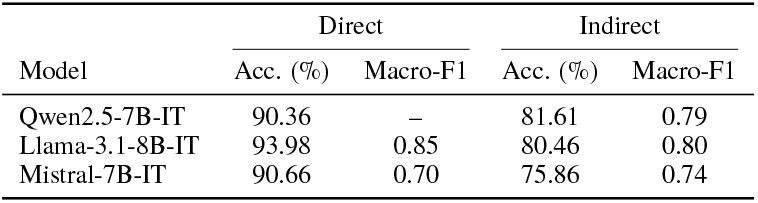
Test set performance of linear probes trained on **C-REACT** (3-way: White/Black/AA/Asian), using Direct vs. Indirect prompt variants.

| Model | Direct |  | Indirect |  |
| --- | --- | --- | --- | --- |
|  | Acc. (%) | Macro-F1 | Acc. (%) | Macro-F1 |
| Qwen2.5-7B-IT | 90.36 | – | 81.61 | 0.79 |
| Llama-3.1-8B-IT | 93.98 | 0.85 | 80.46 | 0.80 |
| Mistral-7B-IT | 90.66 | 0.70 | 75.86 | 0.74 |

For Qwen2.5-7B, which primarily misclassifies White patients as Asian, the Asian Direct neurons show consistently high positive activation regardless of ground truth or prediction. Similarly, Mistral-7B’s tendency toward White → Black/AA errors aligns with elevated activity in Black/AA Direct neurons across most scenarios. Llama-3.1-8B presents a different pattern: while its dominant error is also White → Asian, Asian Indirect neurons show consistently high activation across scenarios. These patterns reveal that neuron groups exhibiting high activation regardless of input correspond to the dominant misclassification directions, suggesting they may act as bias drivers.

#### Intervention Results

Having identified candidate bias drivers, we test whether suppressing these neurons can correct misclassification. Specifically, we evaluate the intervention using three amplification factors (5, 10, 20) to assess if these adjustments alter the model’s predictions. Figure 4 compares correct prediction rates between Direct and Indirect neuron intervention, while Figure 5 shows the full prediction distribution across all conditions.

**Figure 4:**
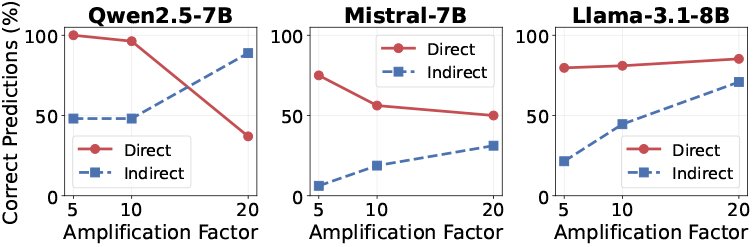
Correct prediction rates after neuron intervention across amplification factors. Direct neuron intervention (solid) generally outperforms Indirect intervention (dashed), demonstrating that neurons encoding explicit racial terminology have stronger causal influence on predictions.

**Figure 5:**
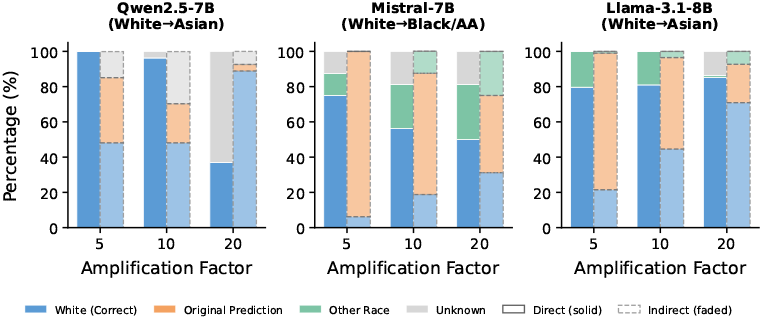
Prediction distribution after neuron intervention on misclassified samples. Direct intervention (solid bars) eliminates the original bias entirely (orange = 0%) across all models and factors, while Indirect intervention (faded bars) leaves residual bias. Higher amplification factors increase Unknown responses (gray)

#### Direct vs. Indirect Neurons

Across all three models, Direct neuron intervention demonstrates stronger causal efficacy than Indirect intervention (Figure 4). At factor 5, Direct intervention achieves substantially higher correct prediction rates across all models compared to Indirect intervention. More importantly, Direct intervention completely eliminates the original bias across all models and amplification factors (Figure 5), while Indirect intervention leaves residual bias. This gap is also consistent with the Llama-3.1-8B pattern: although the Asian Indirect group has higher mean activation in, but higher activation does not necessarily mean stronger causal influence on the final prediction. Indirect cues tend to reflect broad, proxy signals that can be supported by multiple parts of the network, so suppressing one indirect group may be partly compensated elsewhere and leads to a smaller behavioral change. By contrast, Direct neurons are more directly tied to producing the explicit race label, which makes intervening on them more effective.

#### Amplification Factor Selection

The choice of amplification factor involves a tradeoff between bias correction and model stability. We tested factors of 5, 10, and 20, representing increasingly aggressive intervention.

Across all three models, factor 5 yields the best balance. Qwen2.5-7B achieves 100% correct predictions with no Unknown outputs at factor 5, but destabilizes at factor 20 (63% Unknown responses). Mistral-7B reaches 75% correct predictions at factor 5, with higher factors increasingly shifting predictions toward Asian rather than the correct White label. Llama-3.1-8B performs similarly at factors 5 and 10 (approximately 80% correct), with factor 20 introducing Unknown responses. These results suggest that race encoding neurons exert strong influence on predictions, requiring only moderate suppression to alter model behavior. We adopt factor 5 as the default for subsequent analyses.

To illustrate, we provide example model outputs after intervention. Unknown outputs are often malformed, such as “[Yellow] [X] [Black or African” or “[Yellow] The provided options do not include”. In contrast, successful corrections produce valid predictions such as “[White]”.

## 7 Discussion

Our results indicate that the way race and ethnicity are internally represented in LLMs is central to understanding how demographic bias emerges across tasks. The first important finding is that **racial and ethnic concepts are distributed across many internal units rather than localized to a small set of neurons**. Importantly, this distribution is not arbitrary: models decompose race and ethnicity into multiple, interpretable semantic facets, such as explicit group labels and associated geographic or linguistic attributes. Across both ToxiGen and C-REACT, these facets appear as distinct internal representations rather than single abstract concepts (Tables 1, 2, 3). Notably, stereotype-related and historically harmful associations are present across models despite differences in training data and geographic origin, suggesting that bias mitigation cannot rely on a universal map of demographic features but requires model-specific localization.

Secondly, due to this representational structure, the same internal components can be reused across different task contexts, sometimes in ways that lead to biased behavior. **Neurons encoding racial concepts are present in all three models, yet their influence on predictions varies substantially depending on whether the input associates strongly with the proxy cues**. The same representations that benignly encode demographic information can drive biased predictions when activated in contexts where race is irrelevant.

*Crucially, the presence of such representations is not inherently problematic*. Rather, bias arises from how these representations are operationalized during inference. Our intervention *did not erase* racial knowledge from the models; instead, it modulated how this knowledge was reused in task-specific settings. This distinction is critical: pretrained representations reflect what models learn about the world, whereas task-dependent bias reflects when and how those representations are inappropriately applied.

## 8 Conclusion

We provide a mechanistic analysis of how race and ethnicity are represented and operationalized within LLMs. We show that demographic concepts are encoded as distributed, multi-faceted internal representations that can be selectively reused across tasks. These findings suggest that mitigating demographic bias in LLMs requires not only outcome-level interventions, but also a deeper examination of representational structure and task-dependent reuse.

## Data Availability

All data produced are available online at PhysioNet, protected under Data Use Agreement.

## Ethical Statement

This work examine how race and ethnicity are encoded with large language models, which necessarily involves sensitive content including stereotypes and historically offensive terminology. We present these findings to expose potential bias, not to amplify them. We acknowledge that racial categories are socially constructed and vary across cultures; our use of categories reflects the structure of the datasets rather than an endorsement of these taxonomies.

## Acknowledgment

This work was supported by the U.S. National Library of Medicine (NLM), National Institutes of Health, under award number R00LM014308.

## A Appendix

### A.1 Probe Performance

### A.2 Probe Token Projections (ToxiGen)

Table 10 presents the complete top-20 tokens projected by each race direction probe for all three models.

**Table 10:**
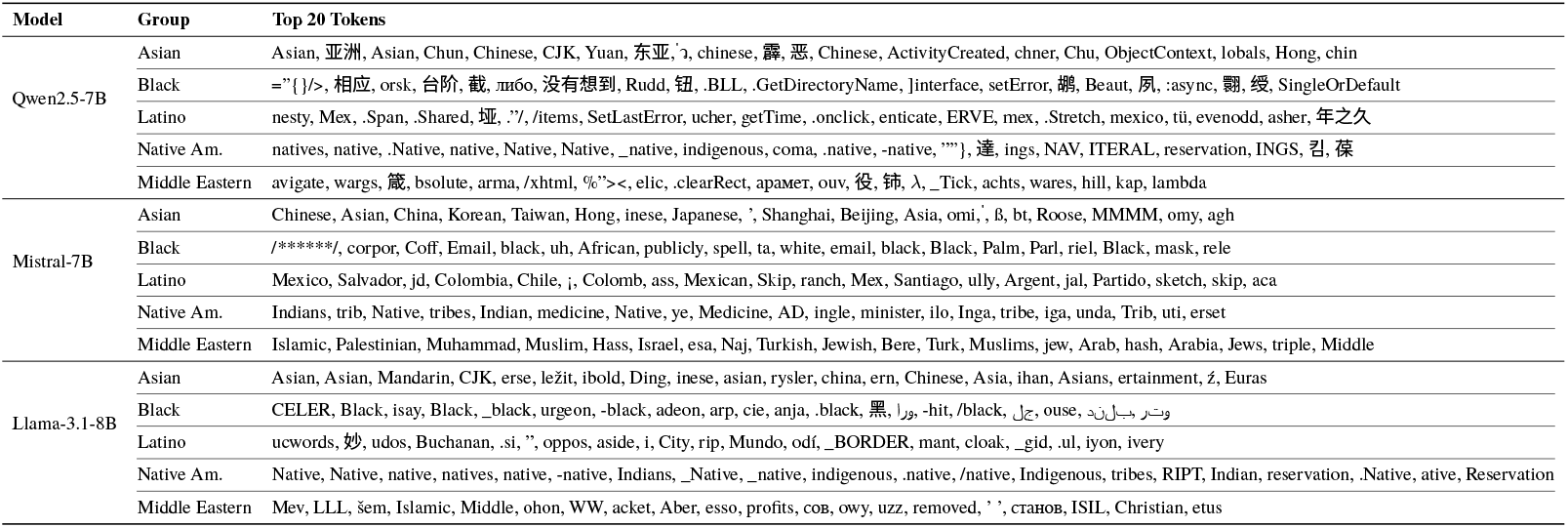
Full top-20 probe token projections for all models (ToxiGen). Tokens are listed in descending order of projection score.

### A.3 Race-Encoding Neurons (ToxiGen)

From Table 11 to Table 13 presents the complete list of race-encoding neurons identified from Toxi-Gen for the three models.

**Table 11:**
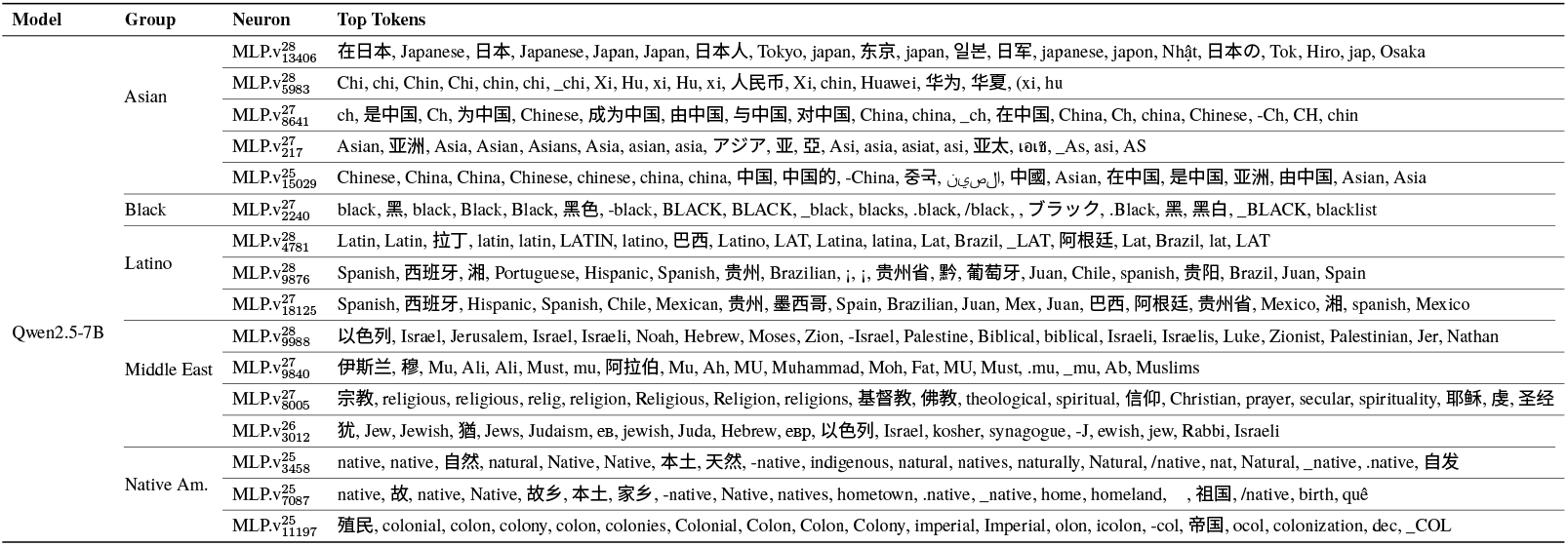
Full top-20 tokens for race-encoding neurons in Qwen2.5-7B (ToxiGen).

**Table 12:**
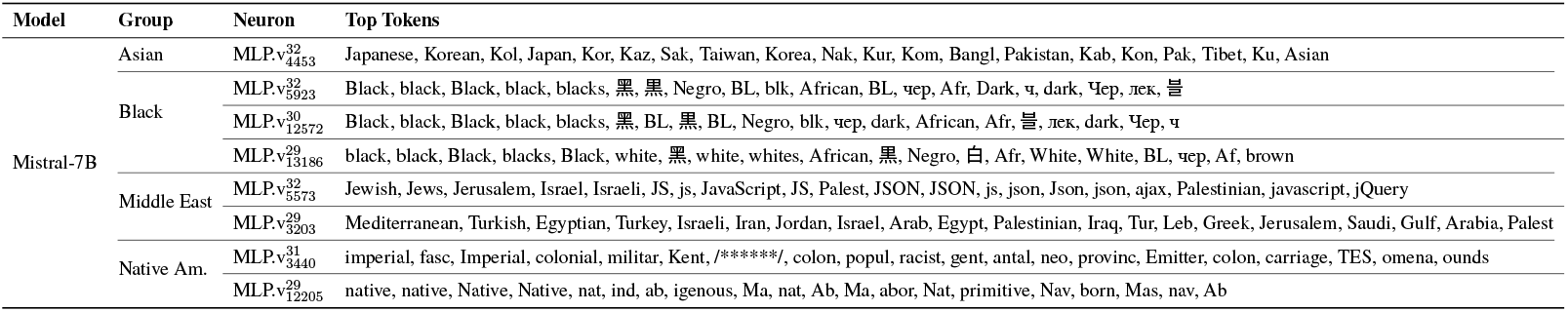
Full top-20 tokens for race-encoding neurons in Mistral-7B (ToxiGen).

**Table 13:**
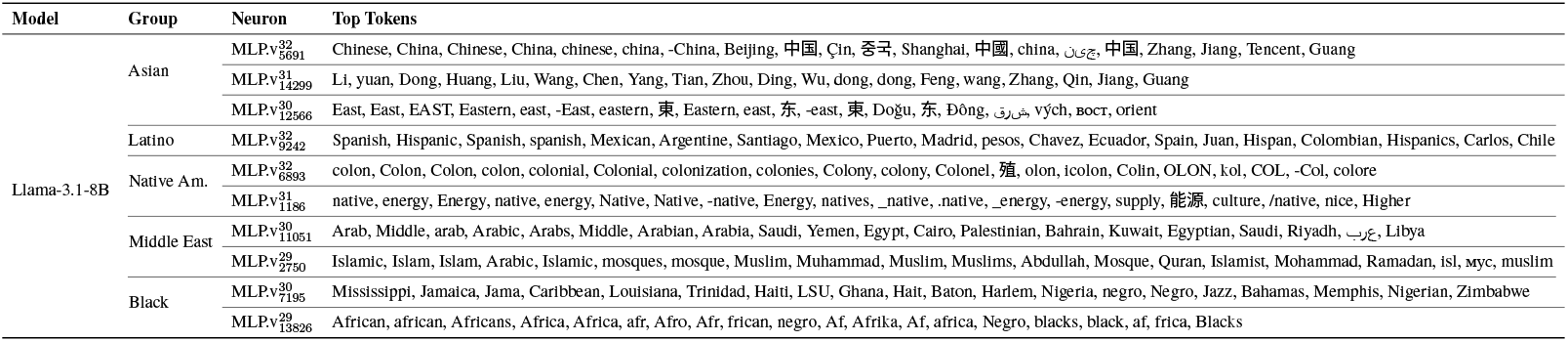
Full top-20 tokens for race-encoding neurons in Llama-3.1-8B (ToxiGen).

### A.4 Probe Token Projections (C-REACT)

Table 14 and Table 15 present the complete top-20 tokens projected by each race direction probe for direct and indirect mentions respectively.

**Table 14:**
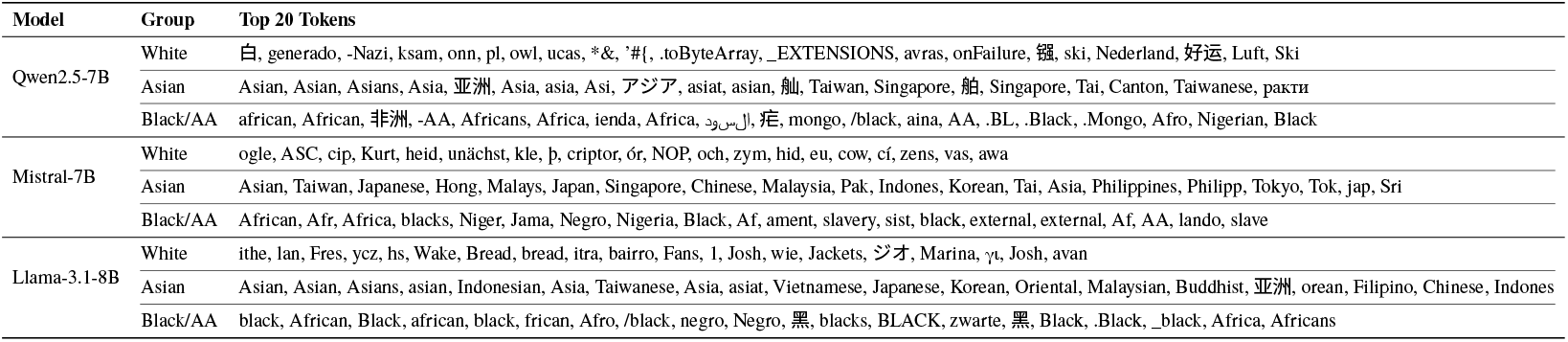
Full top-20 probe token projections for C-REACT direct mentions (explicit race/ethnicity).

**Table 15:**
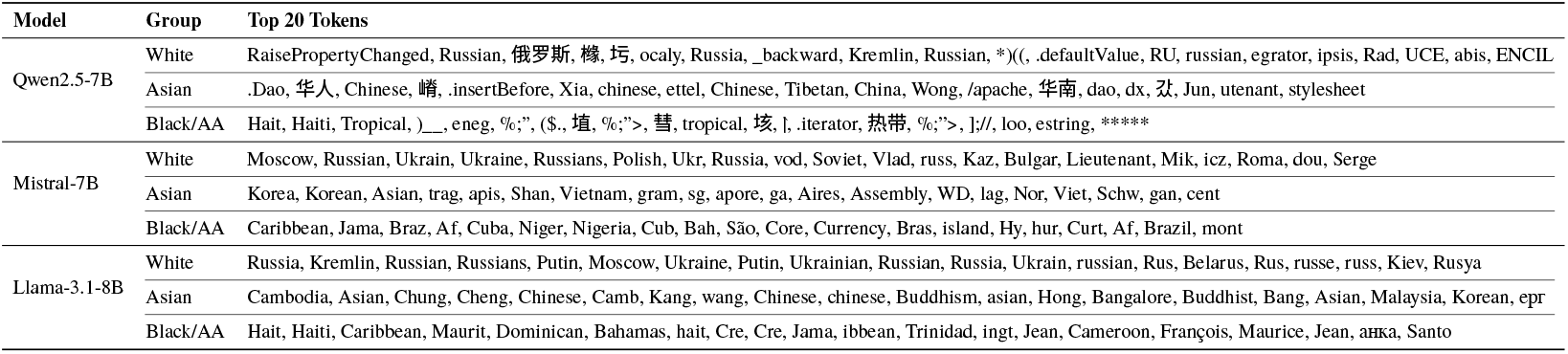
Full top-20 probe token projections for C-REACT indirect mentions (language/country).

| Model | Group | Top 20 Tokens |
| --- | --- | --- |
| Qwen2.5-7B | White | RaisePropertyChanged, Russian, 俄罗斯, 橡, 巧, ocaly, Russia, _backward, Kremlin, Russian, *)((, .defaultValue, RU, russian, egrator, ipsis, Rad, UCE, abis, ENCIL |
|  | Asian | .Dao, 华人, Chinese, 嶙, .insertBefore, Xia, chinese, ettel, Chinese, Tibetan, China, Wong, /apache, 华南, dao, dx, 汎, Jun, utenant, stylesheet |
| | Black/AA | Hait, Haiti, Tropical, )_, eneg, %%;", (\$, 埴, %%;">, 慧, tropical, 垓, ], .iterator, 热带, %%;">, ],/, loo, estring, ***** |
| Mistral-7B | White | Moscow, Russian, Ukrain, Ukraine, Russians, Polish, Ukr, Russia, vod, Soviet, Vlad, russ, Kaz, Bulgar, Lieutenant, Mik, icz, Roma, dou, Serge |
|  | Asian | Korea, Korean, Asian, trag, apis, Shan, Vietnam, gram, sg, apore, ga, Aires, Assembly, WD, lag, Nor, Viet, Schw, gan, cent |
|  | Black/AA | Caribbean, Jama, Braz, Af, Cuba, Niger, Nigeria, Cub, Bah, São, Core, Currency, Bras, island, Hy, hur, Curt, Af, Brazil, mont |
| Llama-3.1-8B | White | Russia, Kremlin, Russian, Russians, Putin, Moscow, Ukraine, Putin, Ukrainian, Russian, Russia, Ukrain, russian, Rus, Belarus, Rus, russe, russ, Kiev, Rusya |
|  | Asian | Cambodia, Asian, Chung, Cheng, Chinese, Camb, Kang, wang, Chinese, chinese, Buddhism, asian, Hong, Bangalore, Buddhist, Bang, Asian, Malaysia, Korean, epr |
|  | Black/AA | Hait, Haiti, Caribbean, Maurit, Dominican, Bahamas, hait, Cre, Cre, Jama, ibbean, Trinidad, ingt, Jean, Cameroon, François, Maurice, Jean, анка, Santo |

### A.5 Race-Encoding Neurons (C-REACT Direct)

Table 16 presents the complete list of race-encoding neurons identified from C-REACT direct mentions (explicit race/ethnicity).

**Table 16:**
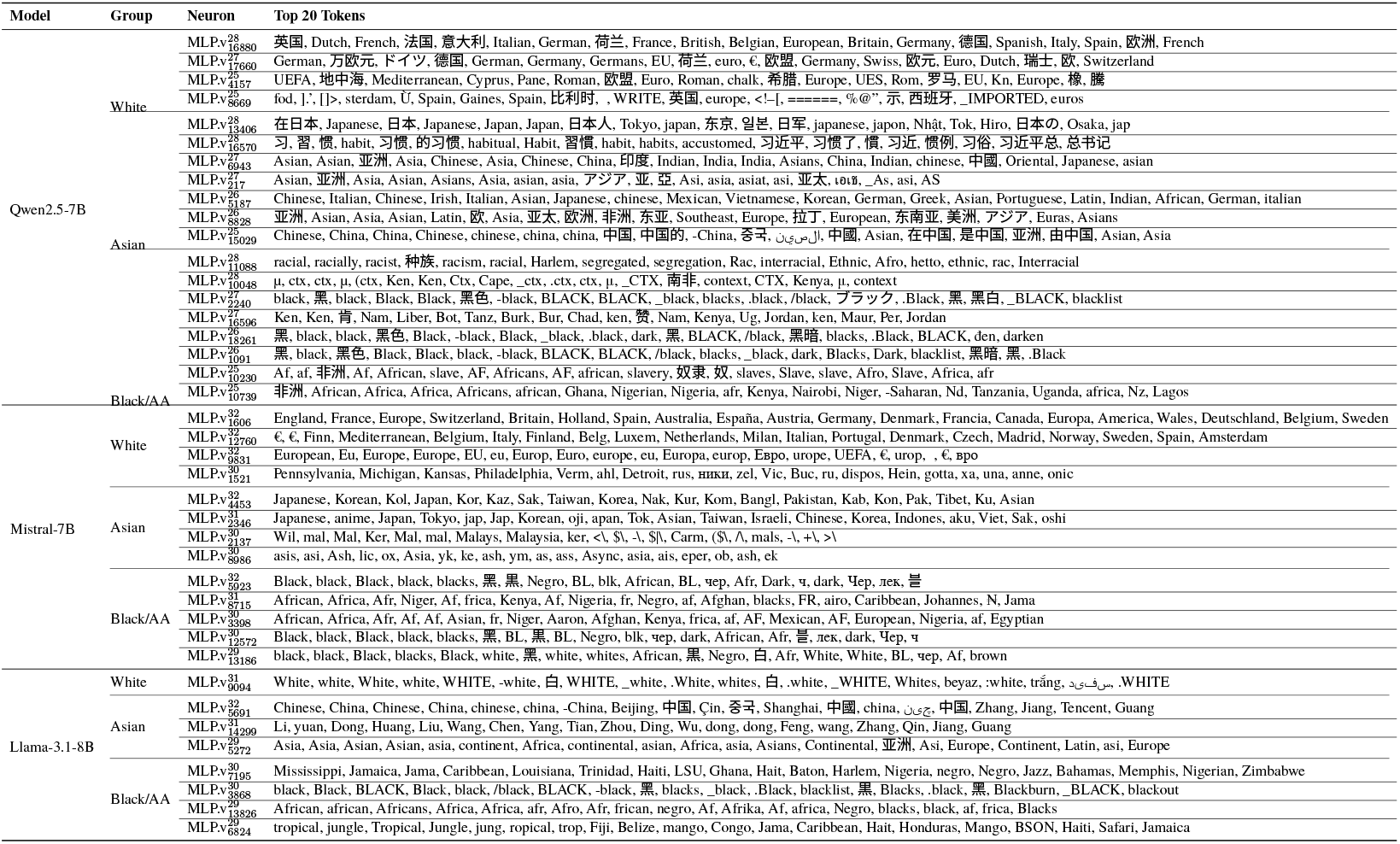
Full list of race-encoding neurons identified from C-REACT direct mentions.

### A.6 Race-Encoding Neurons (C-REACT Indirect)

Table 17 presents the complete list of race-encoding neurons identified from C-REACT indirect mentions (language/country).

**Table 17:**
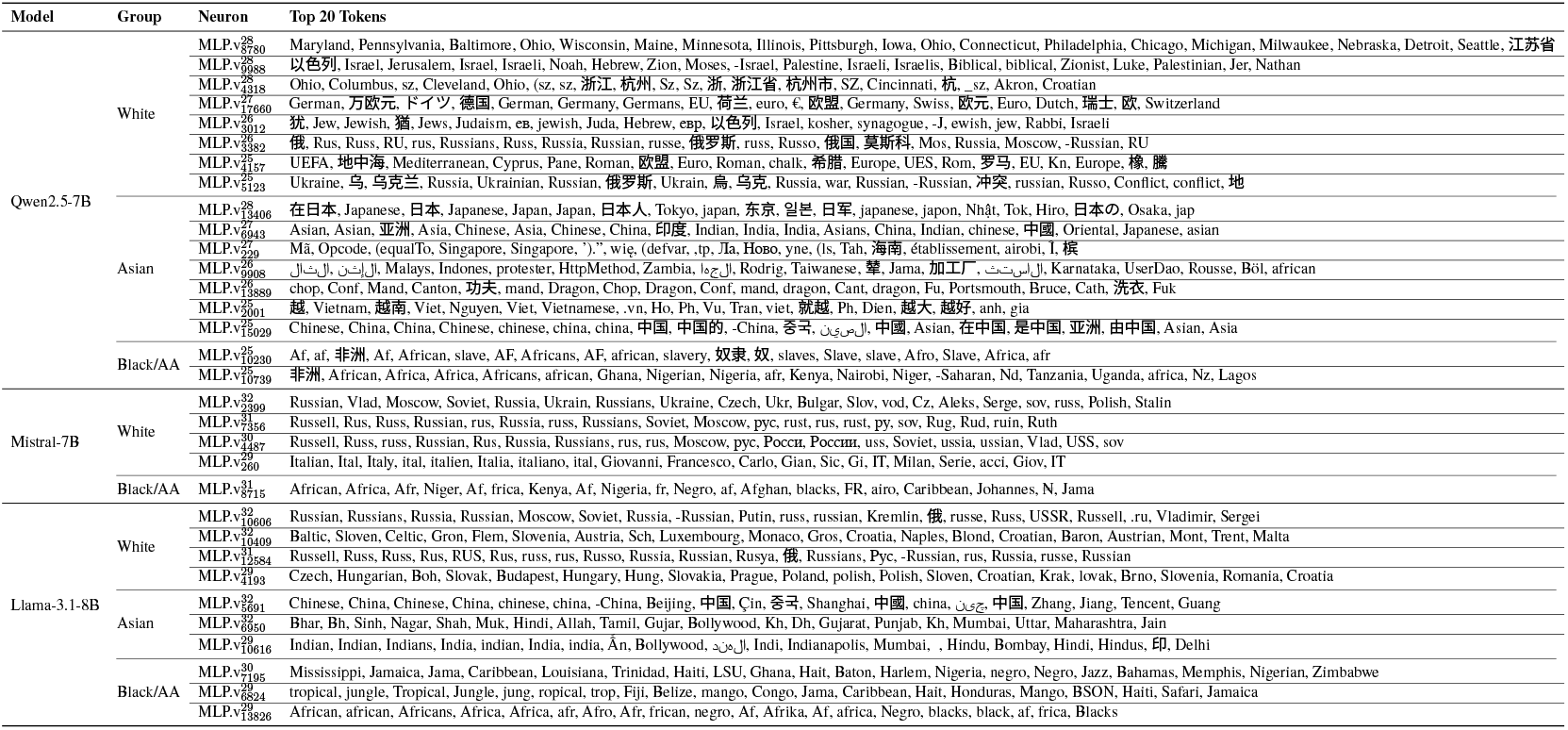
Full list of race-encoding neurons identified from C-REACT indirect mentions (language/country).

## References

Hiba Ahsan, Arnab Sen Sharma, Silvio Amir, David Bau, and Byron C Wallace. 2025. Elucidating mechanisms of demographic bias in llms for healthcare. arXiv preprint 2502.13319.

Hiba Ahsan and Byron C Wallace. 2025. Can saes reveal and mitigate racial biases of llms in healthcare? arXiv preprint 2511.00177.

Oliver Bear Don’t Walk IV, Adrienne Pichon, Harry Reyes Nieva, Tony Sun, Jaan Li, Joshua Winston Joseph, Sivan Kinberg, Lauren R. Richter, Salvatore Crusco, Kyle Kulas, Shaan Ahmed, Daniel Snyder, Ashkon Rahbari, Benjamin Ranard, Pallavi Juneja, Dina Demner-Fushman, and Noemie Elhadad. 2024. C-REACT: Contextualized race and ethnicity annotations for clinical text. PhysioNet. Version 1.0.0.

Nathaniel Demchak, Xin Guan, Zekun Wu, Ziyi Xu, Adriano Koshiyama, and Emre Kazim. 2024. Assessing bias in metric models for llm openended generation bias benchmarks. arXiv preprint 2410.11059.

Kathleen Fraser and Svetlana Kiritchenko. 2024. Examining gender and racial bias in large vision–language models using a novel dataset of parallel images. In Proceedings of the 18th Conference of the European Chapter of the Association for Computational Linguistics (Volume 1: Long Papers), pages 690–713, St. Julian’s, Malta. Association for Computational Linguistics.

Mor Geva, Roei Schuster, Jonathan Berant, and Omer Levy. 2021a. Transformer feed-forward layers are key-value memories. In Proceedings of the 2021 Conference on Empirical Methods in Natural Language Processing, pages 5484–5495.

Mor Geva, Roei Schuster, Jonathan Berant, and Omer Levy. 2021b. Transformer feed-forward layers are key-value memories. In Proceedings of the 2021 Conference on Empirical Methods in Natural Language Processing, pages 5484–5495, Online and Punta Cana, Dominican Republic. Association for Computational Linguistics.

Aaron Grattafiori, Abhimanyu Dubey, Abhinav Jauhri, Abhinav Pandey, Abhishek Kadian, Ahmad Al-Dahle, Aiesha Letman, and et al. 2024. The Llama 3 herd of models. arXiv preprint 2407.21783.

Xin Guan, Nate Demchak, Saloni Gupta, Ze Wang, Ediz Ertekin Jr., Adriano Koshiyama, Emre Kazim, and Zekun Wu. 2025. SAGED: A holistic bias-benchmarking pipeline for language models with customisable fairness calibration. In Proceedings of the 31st International Conference on Computational Linguistics, pages 3002–3026, Abu Dhabi, UAE. Association for Computational Linguistics.

Thomas Hartvigsen, Saadia Gabriel, Hamid Palangi, Maarten Sap, Dipankar Ray, and Ece Kamar. 2022. ToxiGen: A large-scale machine-generated dataset for adversarial and implicit hate speech detection. In Proceedings of the 60th Annual Meeting of the Association for Computational Linguistics (Volume 1: Long Papers), pages 3309–3326, Dublin, Ireland. Association for Computational Linguistics.

Albert Q. Jiang, Alexandre Sablayrolles, Arthur Mensch, Chris Bamford, Devendra Singh Chaplot, Diego de las Casas, Florian Bressand, Gianna Lengyel, Guillaume Lample, Lucile Saulnier, Lélio Renard Lavaud, Marie-Anne Lachaux, Pierre Stock, Teven Le Scao, Thibaut Lavril, Thomas Wang, Timothée Lacroix, and William El Sayed. 2023. Mistral 7B. arXiv preprint 2310.06825.

Adam Karvonen and Samuel Marks. 2025. Robustly improving llm fairness in realistic settings via interpretability. arXiv preprint 2506.10922.

Michelle Kim, Junghwan Kim, and Kristen Johnson. 2023. Race, gender, and age biases in biomedical masked language models. In Findings of the Association for Computational Linguistics: ACL 2023, pages 11806–11815, Toronto, Canada. Association for Computational Linguistics.

Asaf Levartovsky, Mahmud Omar, Girish N Nadkarni, Uri Kopylov, and Eyal Klang. 2025. Sociodemographic bias in large language model– assisted gastroenterology. JAMA Network Open, 8(9):e2532692–e2532692.

Yichen Li, Zhiting Fan, Ruizhe Chen, Xiaotang Gai, Luqi Gong, Yan Zhang, and Zuozhu Liu. 2025. Fairsteer: Inference time debiasing for llms with dynamic activation steering. arXiv preprint 2504.14492.

Afrozah Nadeem, Mark Dras, and Usman Naseem. 2025. Context-aware fairness evaluation and mitigation in llms. arXiv preprint 2510.18914.

nostalgebraist. 2020. interpreting GPT: the logit lens. https://www.lesswrong.com/posts/AcKRB8wDpdaN6v6ru/interpreting-gpt-the-logit-lens. Accessed: 2025-12-23.

Raphael Poulain, Hamed Fayyaz, and Rahmatollah Beheshti. 2024. Bias patterns in the application of llms for clinical decision support: A comprehensive study. arXiv preprint 2404.15149.

Noam Shazeer. 2020. Glu variants improve transformer. arXiv preprint 2002.05202.

Bryan Chen Zhengyu Tan and Roy Ka-Wei Lee. 2025. Unmasking implicit bias: Evaluating persona-prompted LLM responses in power-disparate social scenarios. In Proceedings of the 2025 Conference of the Nations of the Americas Chapter of the Association for Computational Linguistics: Human Language Technologies (Volume 1: Long Papers), pages 1075–1108, Albuquerque, New Mexico. Association for Computational Linguistics.

Qwen Team. 2024. Qwen2.5: A party of foundation models.

Angelina Wang, Michelle Phan, Daniel E. Ho, and Sanmi Koyejo. 2025. Fairness through difference awareness: Measuring Desired group discrimination in LLMs. In Proceedings of the 63rd Annual Meeting of the Association for Computational Linguistics (Volume 1: Long Papers), pages 6867–6893, Vienna, Austria. Association for Computational Linguistics.

Zeping Yu and Sophia Ananiadou. 2025. Understanding and mitigating gender bias in llms via interpretable neuron editing. arXiv preprint 2501.14457.

Travis Zack, Eric Lehman, Mirac Suzgun, Jorge A Rodriguez, Leo Anthony Celi, Judy Gichoya, Dan Jurafsky, Peter Szolovits, David W Bates, Raja-Elie E Abdulnour, and 1 others. 2024. Assessing the potential of gpt-4 to perpetuate racial and gender biases in health care: a model evaluation study. The Lancet Digital Health, 6(1):e12–e22.

Angela Zhang, Mert Yuksekgonul, Joshua Guild, James Zou, and Joseph C Wu. 2023. Chatgpt exhibits gender and racial biases in acute coronary syndrome management. arXiv preprint 2311.14703.

Yachao Zhao, Bo Wang, Yan Wang, Dongming Zhao, Ruifang He, and Yuexian Hou. 2025. Explicit vs. implicit: Investigating social bias in large language models through self-reflection. In Findings of the Association for Computational Linguistics: ACL 2025, pages 1–12, Vienna, Austria. Association for Computational Linguistics.

Hanzhang Zhou, Zijian Feng, Zixiao Zhu, Junlang Qian, and Kezhi Mao. 2024. Unibias: Unveiling and mitigating llm bias through internal attention and ffn manipulation. Advances in Neural Information Processing Systems, 37:102173–102196.

